# Mean corpuscular volume in *HFE* p.C282Y/p.H63D compound heterozygotes with high iron phenotypes: clinical and laboratory associations

**DOI:** 10.1101/2025.10.10.25337747

**Authors:** James C. Barton, J. Clayborn Barton, Ronald T. Acton

**Author notes:** James C. Barton (corresponding author).

## Abstract

**Background:** Variables that influence mean corpuscular volume (MCV) in *HFE* p.C282Y (rs1800562)/p.H63D (rs1799945) compound heterozygotes are inadequately defined.

**Methods:** We retrospectively studied self-reported non-Hispanic white adult compound heterozygotes with transferrin saturation (TS) >50% and serum ferritin (SF) >300 μg/L (men) or TS >45% and SF >200 μg/L (women) who participated in primary care-based screening. In post-screening evaluations, we excluded participants with anemia, pregnancy, or medication use that increases MCV. We defined heavy alcohol intake as >28 g/d men and >14 g/d women. We determined associations of MCV with 11 clinical and laboratory variables.

**Results:** There were 74 participants (37 men, 37 women) of mean age 59±12 (SD) y. Mean screening TS and SF were 65±13% and 529±169 µg/L (men) and 59±14% and 376±195 µg/L (women). Post-screening values did not differ significantly. Mean MCV was 95.7±4.0 fL. There was a negative correlation of MCV with body mass index (p=0.0488) and positive correlations of MCV with age (p=0.0098), daily heme iron intake (p=0.0333), and daily alcohol intake (p=0.0113). Mean MCVs of 19 participants with and 55 without heavy alcohol intake were 97.8±3.8 g/d and 95.0±3.9 g/d, respectively; p=0.0074). Linear regression on MCV confirmed positive associations with age (p=0.0064) and daily alcohol intake (p=0.0151). MCV was not significantly associated with sex, diabetes, daily intakes of non-heme and supplemental iron, swollen or tender 2nd/3rd metacarpophalangeal joints, TS, or SF.

**Conclusion:** MCV in *HFE* p.C282Y/p.H63D compound heterozygotes with high iron phenotypes is positively associated with age and daily alcohol intake, after adjustment for other variables.

## Introduction

*HFE*, the homeostatic iron regulator (chromosome 6p22.2),^1^ ^2^ encodes the cell surface glycoprotein HFE, an upstream regulator of the hepatic hormone hepcidin (*HAMP,* chromosome 19q13.12), the central controller of iron homeostasis.^3^ In persons of European descent, the most common *HFE* missense mutations are p.C282Y (rs1800562) and p.H63D (rs1799945).^4^ The estimated prevalence of p.C282Y/p.H63D compound heterozygotes in non-Hispanic white adults in North America is 1 in 49.^5^ In persons of European descent in the United Kingdom, the estimated prevalence of p.C282Y/p.H63D compound heterozygotes is 1 in 31.^4^

Mean corpuscular volume (MCV) is the average size of red blood cells.^6^ In Western Australia, mean MCVs of *HFE* p.C282Y/p.H63D compound heterozygotes were significantly higher than those of subjects with neither p.C282Y nor p.H63D (*HFE* wt/wt) but were not abnormally elevated.^7^ ^8^ In contrast, the mean MCVs of referred p.C282Y/p.H63D compound heterozygotes in Alabama^9^ and the Netherlands^10^ and corresponding wt/wt control subjects did not differ significantly. In genome wide association studies of adults of European descent, there were positive statistical associations between MCV and p.C282Y^11^ ^12^ ^13^ and p.H63D,^11^ although HFE protein is not expressed in erythroid colonies.^14^ We found no comprehensive study of the relationships between MCV and clinical and laboratory variables in adults with p.C282Y/p.H63D compound heterozygosity.

We performed a retrospective study of 74 adult self-reported non-Hispanic white *HFE* p.C282Y/p.H63D compound heterozygotes who 1) had both elevated TS and elevated SF in a population screening program^15^ and 2) did not have anemia, report pregnancy, or use medications that increase MCV. The aim of this study was to define the relationships between MCV and the following clinical and laboratory variables: age, sex, BMI, diabetes, daily intakes of heme, non-heme, and supplemental iron, daily intakes of alcohol, swollen or tender 2nd/3rd metacarpophalangeal (MCP) joints, TS, and SF. We compare the present observations with those of previously reported hemochromatosis and population cohorts and propose the clinical relevance of the present results.

## Methods

### Ethics approval statement

The Hemochromatosis and Iron Overload Screening (HEIRS) Study, conducted by the National Heart, Lung, and Blood Institute and the National Human Genome Research Institute, in accordance with the principles of the Declaration of Helsinki, evaluated diverse aspects of hemochromatosis, iron overload, and iron-related disorders in a primary care-based sample of 101,168 adults enrolled during the interval 2001-2002 at four Field Centers in the U.S. and one in Canada.^5^ ^15^ ^16^

Local Institutional Review Boards of the HEIRS Study Coordinating Center (Wake Forest University Institutional Review Board, Wake Forest University), the HEIRS Study Central Laboratory (University of Minnesota Institutional Review Board, University of Minnesota), and the HEIRS Study Field Centers (Medical Institutional Review Board, Howard University; UAB Institutional Review Board for Human Use, University of Alabama at Brimingham; University of California Irvine Institutional Review Board, University of California Irvine; Committee for the Protection of Human Subjects/Institutional Review Board, University of Oregon in collaboration with the University of Hawaii Biomedical Institutional Review Board, University of Hawaii/Honolulu; and London Health Sciences Centre Research Institute, London Health Sciences Centre) gave ethical approvals of the Study protocol that is described in detail elsewhere.^5^ ^15^ ^16^

### Participant consent statement

HEIRS Study participants ≥ 25 years of age were recruited from outpatient facilities affiliated with the Field Centers and gave written informed consent for screening and post-screening evaluation.^5^ ^15^ ^16^ The HEIRS Study informed consent forms, not available as public documents, were used during the participant recruitment phase of the Study (2001-2002). Each of the Field Centers in North America used an Institutional Review Board-approved consent form tailored to its specific institution.^5^ ^15^ ^16^

### Primary care-based screening

The HEIRS Study recruited participants from public and private primary care offices in ambulatory clinics, a health maintenance organization, and diagnostic blood collection centers affiliated with five Field Centers.^5^ Ninety-eight percent of self-reported non-Hispanic white participants were recruited at Field Centers in Alabama, California, Ontario, and Oregon/Hawaii.^17^ Laboratory testing at screening included only TS and SF phenotyping and *HFE* p.C282Y and p.H63D allele-specific genotyping.^5^ p.C282Y/p.H63D compound heterozygosity was identified in 908 of 44,082 non-Hispanic white adult screening participants (2.06% [95% confidence interval (CI): 1.93, 2.91]).^17^ Medical histories were not compiled and physical examinations were not performed at primary care-based screening.

### Post-screening evaluation attendees

Invitations to attend post-screening evaluations were extended to all 78 participants with *HFE* p.C282Y/p.H63D compound heterozygosity whose screening TS and SF values were elevated (TS >50% and SF >300 μg/L for men; TS >45% and SF >200 μg/L for women).^15^ ^18^ Post-screening evaluation attendees included 77 of the 78 invitees (39 men, 38 women). We excluded two men and one woman because they had anemia (hemoglobin (Hb) <130 g/L (men) and <120 g/L (women), pregnancy, or medication use that increased MCV.^19^ The cohort for analysis consisted of 74 participants (37 men, 37 women).

### Post-screening evaluations

Evaluations included the following: 1) questionnaires completed by participants that addressed medical histories and medications;^18^ 2) University of Hawaii Multiethnic Dietary Questionnaires;^18^ ^20^ 3) focused physical examinations performed by HEIRS Study physicians;^18^ and 4) laboratory testing of blood specimens.^18^ The median interval between primary care-based screening and post-screening evaluations was eight months.^18^

### Medical history questionnaires

We defined reports of diabetes as affirmative responses to this question: “Have you ever been told you have diabetes?” We defined reports of cirrhosis as affirmative responses to this question: “Ever told that you have/had cirrhosis?”

### Dietary questionnaires

Analyses of Multiethnic Dietary Questionnaires at the University of Hawaii provided estimates of the average daily intakes of dietary iron, supplemental iron, and alcohol for the previous year.^18^ ^20^ The dietary iron attributed to intakes of meat, fish, and poultry was classified as heme iron and other dietary iron as non-heme iron. Iron intakes were expressed as mg/d. Alcohol intakes were expressed as g/d. We defined heavy alcohol intake as the average intake of more than 14 standard drinks per week (> 28 g/d) for men and more than 7 standard drinks per week (> 14 g/d) for women.^21^

### Physical examinations

We used Quetelet’s formula (kg/m^2^) to measure body mass index (BMI).^22^ Physicians recorded the presence or absence of swelling or tenderness of the 2nd/3rd metacarpophalangeal (MCP) joints.^18^

### Laboratory testing

Blood samples for post-screening testing were obtained after an overnight fast. All participants were tested for *HFE* genotype confirmation, complete blood count, TS, and SF.^5^ Serum alanine aminotransferase (ALT) and aspartate aminotransferase (AST) levels were available in 46 participants (62.2%). All testing was performed at the HEIRS Study Central Laboratory (Fairview-University Medical Center Clinical Laboratory, University of Minnesota, Fairview, MN, USA).^15^ ^18^

### Markers of advanced hepatic fibrosis or cirrhosis

We calculated fibrosis-4 (FIB-4) indices^23^ and AST-to-platelet ratio indices (APRI)^24^ as markers of advanced hepatic fibrosis or cirrhosis risk in the 46 participants (62.2%; 25 men, 21 women) for whom the requisite data were available. We classified risks with FIB-4 as: low (FIB-4 < 1.3); intermediate (FIB-4 1.3 - 2.67); and high (FIB-4 > 2.67).^23^ We classified risks with APRI as: low (APRI < 0.05); intermediate (APRI 0.5 - 1.5); and high (APRI > 1.5).^24^ We also compiled participant reports of cirrhosis diagnoses.

### Statistics

The dataset for analyses consisted of complete observations on 74 participants (37 men, 37 women) except FIB-4 and APRI indices (25 men, 21 women). Kolmogorov-Smirnov testing demonstrated that age, BMI, daily intake of heme iron, TS, SF, and MCV data did not differ significantly from those that are normally distributed. We displayed these data as means ± 1 standard deviation (SD) and compared means using the Student’s t test for unpaired samples (two-tailed). We displayed other continuous data as medians (ranges) and compared medians using the Mann-Whitney U test (two-tailed). We compared binary data using Fisher’s exact test (two-tailed). We computed associations of MCV with available continuous variables using Pearson’s correlation (two-tailed) or Spearman’s rank correlation (two-tailed) for paired data, as appropriate.

We evaluated these 11 independent variables for suitability in a multiple linear regression on MCV: age, BMI, sex, diabetes, intakes of heme, non-heme, and supplemental iron and alcohol, swollen or tender 2nd/3rd MCP joints, TS, and SF. We deleted sex because it was significantly associated with SF in a Bonferroni-corrected correlation matrix. A preliminary regression on MCV using the remaining variables revealed low standardized beta coefficients (beta) and high values of p for BMI, diabetes, non-heme iron intake, swollen or tender 2nd/3rd MCP joints, TS, and SF. Thus, we excluded these independent variables. We report the contributions of significant remaining independent variables to the final regression as beta and the proportion of variance in MCV explained by the independent variable(s) as R².

We used Excel^®^ 2022 (Microsoft Corp., Redmond, WA, USA) and GraphPad Prism 8^®^ (2018; GraphPad Software, San Diego, CA, USA). We defined p <0.05 to be significant.

## Results

### Characteristics of 74 *HFE* p.C282Y/p.H63D compound heterozygotes

There were 37 men and 37 women of mean age of 59 ± 12 y. Respective mean TS and mean SF values in men and women in screening and post-screening evaluations did not differ significantly (Table 1). Mean MCV was 95.7 ± 4.0 fL. Mean SF and Hb were higher in men (Table 2). Nine participants (12.2%; 4 men, 5 women) had MCV ≥ 100.0 fL.

**Table 1.** Iron phenotypes in 74 *HFE* p.C282Y/p.H63D compound heterozygotes^a^.

| Men (n = 37) | Screening | Post-screening evaluation | Value of p |
| --- | --- | --- | --- |
| Mean transferrin saturation, % ± 1 SD | $65 \pm 13$ | $58 \pm 17$ | 0.0692 |
| Mean serum ferritin, $\mu\text{g/L} \pm 1$ SD | $529 \pm 169$ | $531 \pm 170$ | 0.9684 |
| Women (n = 37) |  |  |  |
| Mean transferrin saturation, % ± 1 SD | $59 \pm 14$ | $53 \pm 12$ | 0.0594 |
| Mean serum ferritin, $\mu\text{g/L} \pm 1$ SD | $376 \pm 195$ | $366 \pm 201$ | 0.8316 |

<sup>a</sup> SD, standard deviation

**Table 2.** *HFE* p.C282Y/p.H63D compound heterozygotes in post-screening evaluations^a^.

| Variable | Men (n = 37) | Women (n = 37) | Value of p |
| --- | --- | --- | --- |
| Mean age, y (SD) | $57 \pm 13$ | $62 \pm 12$ | 0.0606 |
| Mean body mass index, $\text{kg/m}^2$ (SD) | $28.7 \pm 5.0$ | $28.0 \pm 5.2$ | 0.5405 |
| Diabetes reports, % (n) | 24.3 (6) | 5.4 (2) | 0.2611 |
| Mean heme iron intake, mg/d (SD) | $2.4 \pm 1.4$ | $1.9 \pm 1.0$ | 0.0549 |
| Median non-heme iron intake, mg/d (range) | 12.3 (4.2, 34.9) | 12.2 (4.3, 43.7) | 0.4822 |
| Median supplemental iron intake, mg/d (range) | 3.9 (0, 40.5) | 3.9 (0, 40.5) | 0.6892 |
| Median alcohol intake, g/d (range) | 5.7 (0, 60.9) | 5.7 (0, 55.8) | 0.7949 |
| Swollen or tender 2nd/3rd MCP joints, % (n) | 5.4 (2) | 5.4 (2) | ~1.0000 |
| Mean transferrin saturation, % (SD) | $58 \pm 17$ | $53 \pm 12$ | 0.1197 |
| Mean serum ferritin, $\mu\text{g/L}$ (SD) | $531 \pm 170$ | $366 \pm 201$ | 0.0003 |
| Mean hemoglobin, g/L | $155 \pm 8$ | $139 \pm 9$ | < 0.0001 |
| Mean corpuscular volume, fL (SD) | $95.5 \pm 4.0$ | $96.0 \pm 4.1$ fL | 0.6087 |

<sup>a</sup> MCP, metacarpophalangeal; SD, standard deviation

### Markers of advanced hepatic fibrosis

High-risk FIB-4 indices were observed in five of 46 participants (10.9%), none of whom had a high-risk APRI (Table 3). There was no significant difference in mean MCV by low, intermediate, or high risk FIB-4 or APRI classification (Table 3). One woman reported that she was diagnosed with cirrhosis, although both her FIB-4 and APRI risks were classified as intermediate.

**Table 3.** Hepatic fibrosis risk in 46 *HFE* p.C282Y/p.H63D compound heterozygotes^a^.

|  | Low risk, % (n) | Intermediate risk, % (n) | High risk, % (n) | Value of p |
| --- | --- | --- | --- | --- |
| FIB-4<sup>b</sup> | 56.5 (26) | 32.6 (15) | 10.9 (5) | - |
| Mean MCV, fL (SD) | 95.7 ± 3.5 | 96.3 ± 5.6 | 97.0 ± 5.1 | 0.8204<sup>d</sup> |
| APRI<sup>c</sup> | 78.3 (36) | 21.7 (10) | 0 (0) | - |
| Mean MCV, fL (SD) | 96.4 ± 4.0 | 94.8 ± 5.5 | - | 0.3140<sup>e</sup> |

<sup>a</sup> APRI, AST-to-platelet ratio index; FIB-4, fibrosis-4 index; MCV, mean corpuscular volume; SD, standard deviation
<sup>b</sup> low risk (FIB-4 < 1.3); intermediate risk (FIB-4 1.3 - 2.67); and high risk (FIB-4 > 2.67).
<sup>c</sup> low risk (APRI < 0.5); intermediate risk (APRI 0.5 - 1.5); and high risk (APRI > 1.5).
<sup>d</sup> One-way ANOVA p test.
<sup>e</sup> Unpaired Student t test (two-tailed).

### MCV correlations with continuous variables

There were significant positive correlations of MCV with age (Fig. 1, Table 4), daily heme iron intake (Table 4), and daily alcohol intake (Fig. 2, Table 4), and a significant negative correlation of MCV with BMI (Table 4).

**Figure 1.**
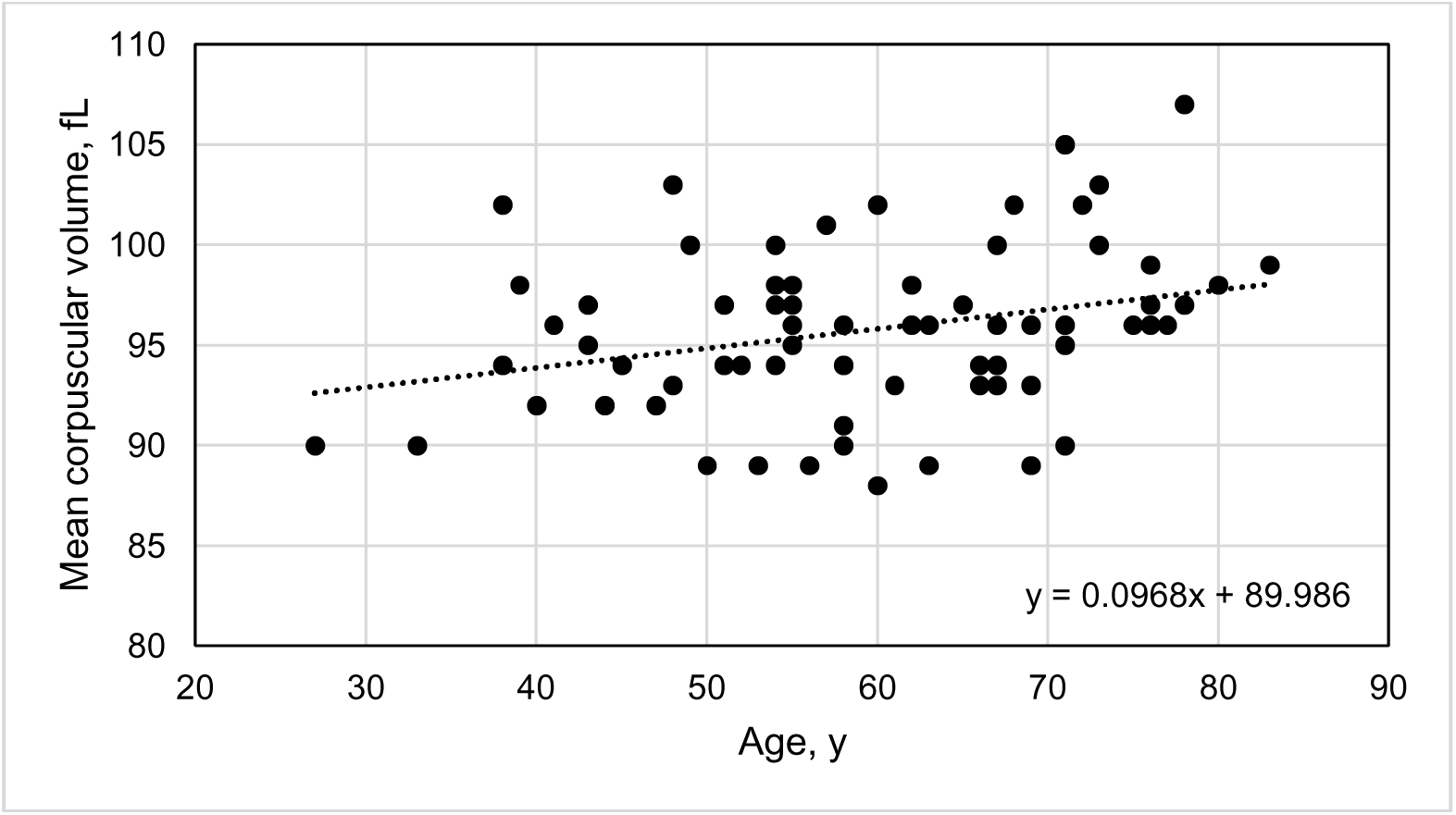
Pearson’s correlation of mean corpuscular volume vs. age in 74 *HFE* p.C282Y/p.H63D compound heterozygotes with high iron phenotypes (r_74_ = 0.2983; p = 0.0098).

**Figure 2.**
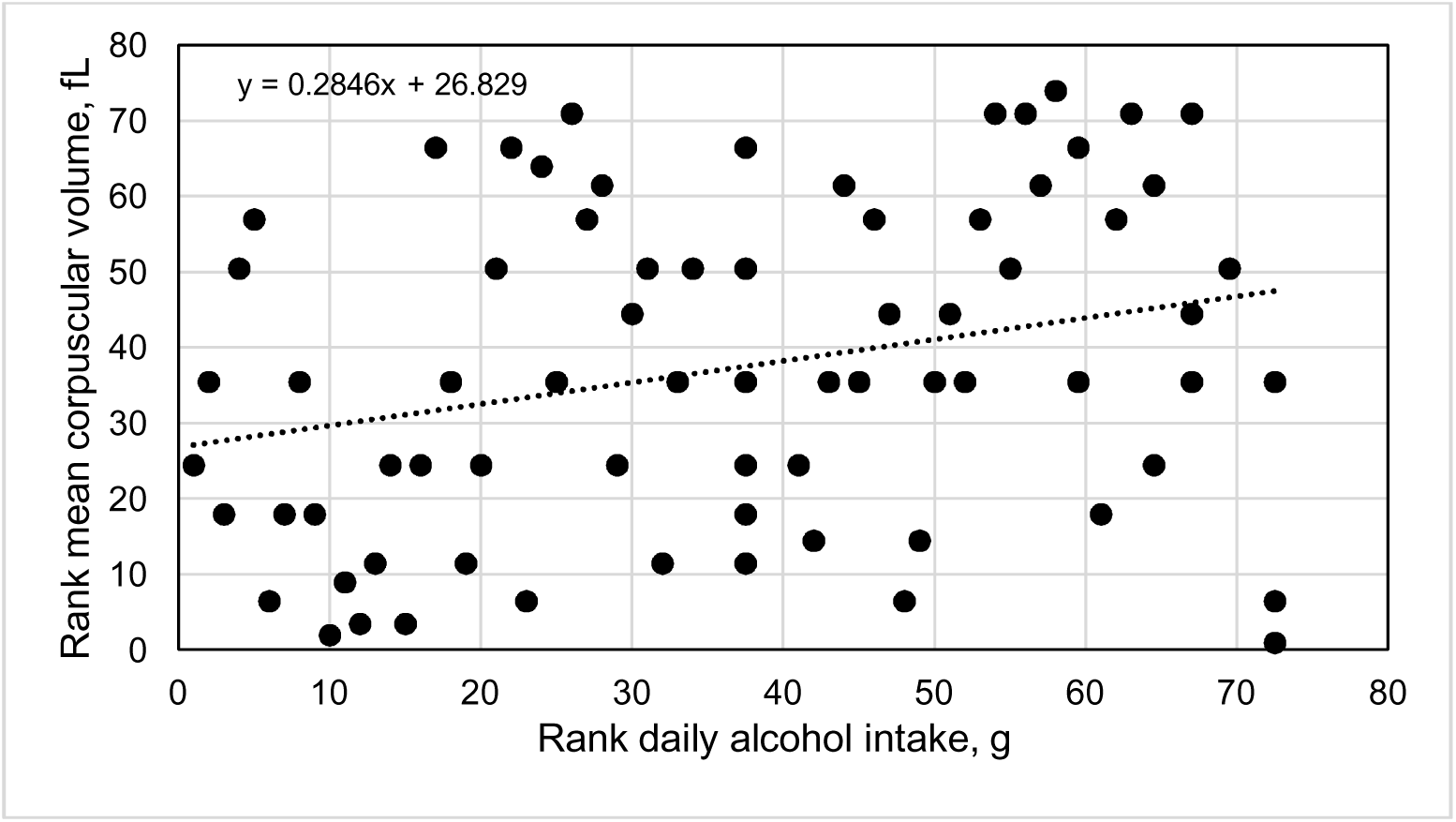
Spearman’s rank correlation of mean corpuscular volume vs. daily alcohol intake in 74 *HFE* p.C282Y/p.H63D compound heterozygotes with high iron phenotypes (*r*_74_ = 0.2860; p = 0.0135).

**Table 4.** Correlations of mean corpuscular volume in 74 *HFE* p.C282Y/p.H63D compound heterozygotes.

| Variable | Pearson's coefficient *r* | Spearman's coefficient *r* | Value of *p*<sup>a</sup> |
| --- | --- | --- | --- |
| Age, y | 0.2983 | - | 0.0098<sup>a</sup> |
| Body mass index, kg/m<sup>2</sup> | -0.2299 | - | 0.0488 |
| Heme iron intake, mg/d | -0.2478 | - | 0.0333 |
| Non-heme iron intake, mg/d | - | -0.0604 | 0.6092 |
| Supplemental iron intake, mg/d | - | 0.0742 | 0.5300 |
| Alcohol intake, g/d | - | 0.2860 | 0.0135<sup>b</sup> |
| Transferrin saturation, % | 0.0667 | - | 0.5723 |
| Serum ferritin, µg/L | -0.0403 | - | 0.7296 |

<sup>a</sup> See Figure 1.
<sup>b</sup> See Figure 2.

### Mean MCV and binary variables

Mean MCV did not differ significantly between men and women or between participants with and without diabetes reports or swollen or tender 2nd/3rd MCP joints (Table 5).

**Table 5.** Mean corpuscular volume in 74 *HFE* p.C282Y/p.H63D compound heterozygotes^a^.

| Variable | Mean MCV, fL ± SD with variable (n) | Mean MCV, fL ± SD without variable (n) | Value of p |
| --- | --- | --- | --- |
| Male sex | 95.5 ± 4.0 (37) | 96.0 ± 4.1 (37) | 0.6087 |
| Diabetes reports | 94.6 ± 3.3 (8) | 95.8 ± 4.2 (66) | 0.4045 |
| Swollen or tender 2nd/3rd MCP joints | 97.0 ± 5.5 (4) | 95.6 ± 4.0 (70) | 0.6768 |

<sup>a</sup> MCP, metacarpophalangeal; MCV, mean corpuscular volume; SD, standard deviation

### MCV and heavy alcohol intakes

The mean MCV of the 19 participants (10 men, 9 women) with heavy alcohol intakes was greater than the mean MCV of the other 55 participants (97.8 ± 3.8 vs. 95.0 ± 3.9, respectively; p = 0.0074). Four men and two women with heavy alcohol intakes had MCV ≥ 100 fL.

### Regression on MCV

Multiple linear regression on MCV using the independent variables age, daily heme iron intake, and daily alcohol intake revealed positive associations with age (p = 0.0064; beta 0.3055) and daily alcohol intake (p = 0.0151; beta 0.2707). The R^2^ (adjusted R^2^) was 0.1622 (0.1386). ANOVA p of this regression was 0.0019.

## Discussion

The novel findings of this study of 74 *HFE* p.C282Y/p.H63D compound heterozygotes with elevated screening TS and SF are that MCV was significantly associated with both age and daily alcohol intake after adjustment for other variables. An unexpected finding is that MCV was not significantly associated with TS, in contrast to observations in p.C282Y homozygotes.^9^ ^25^ ^26^ ^27^

Increasing MCV of the present *HFE* p.C282Y/p.H63D compound heterozygotes was associated with increasing age. MCV also increased with age in post-screening p.C282Y homozygotes^27^ and English adults in primary care venues.^28^ Increasing age^29^ and MCV^30^ are inversely related to leukocyte telomere lengths (LTLs). In a Mendelian randomization study, an increase of one standard deviation in genetically influenced telomere length decreased MCV significantly.^31^ In another study, mean LTL in p.C282Y homozygotes with and without elevated iron phenotypes did not differ significantly.^32^ These observations suggest that genetic determinants of telomere length,^31^ not *HFE* genotypes or iron phenotypes, account in part for the age-related increase in MCV we observed in the present p.C282Y/p.H63D compound heterozygotes.

There was a moderate negative correlation of MCV with BMI in this study, although MCV was not significantly associated with BMI after adjustment for other variables. In self-reported healthy subjects aged > 18 y in the 2011-2016 National Health and Nutrition Examination Survey, there was a weak negative association of MCV with BMI.^33^

Mean MCV of the present *HFE* p.C282Y/p.H63D compound heterozygotes with and without diabetes reports did not differ significantly. The mean MCV of Scottish adults with diabetes was significantly higher than that of control subjects.^34^ In contrast, the mean MCV of Indian adults with type 2 diabetes was significantly lower than that of control subjects.^35^

There was a weak positive correlation of MCV with daily heme iron intake in this study, although MCV was not significantly associated with daily heme iron intake after adjustment for other variables. In Dutch blood and plasma donors, MCV was positively associated with heme iron intakes in men but not in women.^36^

MCVs of the present *HFE* p.C282Y/p.H63D compound heterozygotes were positively associated with daily alcohol intakes. The mean MCV of Australian men and women with any p.C282Y genotype also increased with alcohol intake after adjustment for age.^37^ In the same study, neither p.C282Y nor p.H63D alone predisposed to moderate or heavy alcohol intake.^37^ In British adults without anemia, MCV also increased with alcohol intake, but there was no interaction between alcohol consumption and allelic variants associated with MCV.^38^

The present *HFE* p.C282Y/p.H63D compound heterozygotes were invited to attend post-screening evaluations because their screening TS and SF values were elevated.^15^ Heavy alcohol intake occurred in 25.7% of the present compound heterozygotes, whereas heavy alcohol intake occurs in ∼6.5% of non-Hispanic white adults in the U.S.^21^ Alcohol intake was excessive in 38.9% of Australian p.C282Y/p.H63D compound heterozygotes with hemochromatosis and SF > 1000 µg/L.^39^ Excessive alcohol intake increases TS^40^ ^41^ and SF.^42^ The clinical significance of these observations is that alcohol intake is significantly associated with elevated MCV in p.C282Y/p.H63D compound heterozygotes with both elevated TS and elevated SF.

Swollen or tender 2nd/3rd MCP joints were not associated with MCV and macrocytosis in this study, in contrast to observations in *HFE* p.C282Y homozygotes.^27^ ^43^ The prevalence of swollen or tender 2nd/3rd MCP joints in the present p.C282Y/p.H63D compound heterozygotes was lower than that of post-screening participants with p.C282Y homozygosity (5.4% and 14.4%, respectively; p = 0.0246).^27^

It was unexpected that MCVs in the present *HFE* p.C282Y/p.H63D compound heterozygotes were not significantly associated with TS, in contrast to the positive associations between MCV and TS in p.C282Y homozygotes.^9^ ^26^ ^27^ Mean TS is significantly lower in untreated p.C282Y/p.H63D compound heterozygotes than untreated p.C282Y homozygotes.^5^ This suggests that the mean TS in p.C282Y/p.H63D compound heterozygotes is insufficient to increase the uptake of transferrin-bound iron by immature erythroid cells, increase Hb synthesis, and thereby increase MCV.^9^

Limitations of this study include the lack of observations of *HFE* p.C282Y/p.H63D participants without both elevated TS and SF and of participants aged < 25 y.^5^ The HEIRS Study did not measure serum levels of vitamin B12 or folate, although none of the present subjects had anemia and deficiencies of these micronutrients were uncommon causes of macrocytosis in another primary care cohort.^44^ More than three-quarters of the variance in MCV values in the present linear regression is attributable to variables we did not analyze, especially non-*HFE* genetic factors.^11^ ^31^ ^38^ Measuring iron stores using quantitative phlebotomy or liver specimens obtained by biopsy and performing clinical evaluations for cirrhosis were beyond the scope of the cross-sectional HEIRS Study.

The present post-screening examination invitation criteria of both elevated TS and elevated SF may have preferentially although unintentionally selected p.C282Y/p.H63D compound heterozygotes with heavy daily alcohol intakes. None of 46 participants had combination of high-risk FIB-4 and APRI indices. It is unknown whether any of the other 28 participants (37.8%) would have had high-risk FIB-4 and APRI indices or not, although none of them reported a diagnosis of cirrhosis. The prevalence of documented iron overload-related disease (including cirrhosis) was low in other cohorts of *HFE* p.C282Y/p.H63D compound heterozygotes.^45^ ^46^ These observations suggest the prevalence of occult advanced hepatic fibrosis or cirrhosis is also low in the present cohort and unlikely to influence the present results significantly.

## Conclusion

We conclude that MCV in *HFE* p.C282Y/p.H63D compound heterozygotes with high iron phenotypes is positively associated with age and daily alcohol intake, after adjustment for other variables.

## Data Availability

The National Heart, Lung, and Blood Institute provides controlled access to individual participant data through the Biologic Specimen and Data Repository Information Coordinating Center (BioLINCC) (https://biolincc.nhlbi.nih.gov/studies/heirs/). Data access requires registration, evidence of local institutional review board approval or certification of exemption from institutional review board review, and completion of a data use agreement. The National Heart, Lung, and Blood Institute does not permit investigators to submit data directly to journals, related repositories, or other sources. Parties interested in obtaining the data analyzed in the present study are referred to BioLINCC.

https://pubmed.ncbi.nlm.nih.gov/12589228/

## Acknowledgements

The following individuals also performed data collection (2001-2003) and received compensation for their work: Paul C. Adams, MD (Department of Medicine, London Health Sciences Centre, London, Ontario, Canada); John H. Eckfeldt, MD, PhD (Department of Laboratory Medicine and Pathology, University of Minnesota, Minneapolis); Victor R. Gordeuk, MD (Division of Hematology and Oncology, Department of Medicine, University of Illinois at Chicago, Chicago); Emily Harris, PhD (Epidemiology and Genomics Research Program, Division of Cancer Control and Population Sciences, National Cancer Institute, National Institutes of Health, Bethesda, Maryland); Helen Harrison, RN (The Western-Fanshawe Collaborative BScN Program, Fanshawe College, London, Ontario, Canada); Christine E. McLaren, PhD (Department of Epidemiology, University of California, Irvine, California); and Gordon D. McLaren, MD (Division of Hematology/Oncology, Department of Medicine, University of California, Irvine, and Department of Veterans Affairs Long Beach Healthcare System, Long Beach, California).

## Authors’ contributions

Each author contributed substantively to this study. JaCB conceived this study and its methodology, collected data, evaluated participants in post-screening evaluations, performed analyses, and drafted the manuscript. JClB conceived study methodology, curated data, performed analyses, and drafted the manuscript. RTA conceived this study and its methodology, collected data, performed analyses, curated data, and drafted the manuscript. Each author approved the manuscript in its final form.

